# An easy, reliable and rapid SARS-CoV2 RT-LAMP based test for Point-of-Care and diagnostic lab

**DOI:** 10.1101/2020.09.25.20200956

**Authors:** Meriadeg AR Gouilh, Renaud Cassier, Elodie Maille, Cécile Schanen, Louis-Marie Rocque, Astrid Vabret

## Abstract

This study presents and evaluates a rapid and all-in-one SARS-CoV-2 RT-LAMP based molecular detection system, including RNA extraction or not, for point-of-care or massive testing of naso-pharyngeal swabs. The point-of-care format uses LoopX©, a small portative device ensuring optimal LAMP reaction and automated reading with 95.2% and 95.5% sensitivity and specificity respectively. This system might also be useful for testing other sample types such as saliva.

## CONTEXT

In late December 2019, a new disease named COVID-19 (COronaVirus Infectious Disease 2019), due to a new *Sarbecovirus, Betacoronavirus*, the SARS-CoV-2, was notified in Wuhan, China. WHO declared “A public health emergency of international concern” on January 30th 2020. Six months later, the virus has spread to the whole world and has caused more than 14 000,000 cases including more than 616,000 deaths (with a large majority in USA and Europe - 22 July 2020, WHO).

The ability of both accurate and massive testing is paramount for adequate response to viral diffusion by the implementation of public health control measures. Nasopharyngeal swab (NPS) is the most widely used and the most reliable method of sampling for respiratory viruses detection. To date, NPS collected in liquid universal transport media and combined with standardised nucleic-acid extraction and real time RT-PCR (RT-qPCR), is the gold standard method to detect SARS-CoV-2. Nevertheless, these steps still necessitate from 4 to 8 hours of handling and incubation times which in turn induces a sample-to-result time ranging from 6 to 24 hours in diagnostic labs. These time constraints, processing delays and the necessity of expert technicians remain a major impediment to the reactivity and efficiency of diagnostic laboratories and to testing policies.

## RT-LAMP

(Reverse-Transcription and Loop-mediated isothermal AMPlification) might advantageously increase testing efficiency as it allows all-in-one rapid test including RNA reverse transcription and DNA amplification without temperature cycles requirement. LAMP does not even require complex devices and laboratory infrastructures, RT-LAMP can use low-cost and mobile equipment already available in most molecular diagnostic laboratories and utilises the inhibitors tolerant Bst polymerase [1]. These traits make LAMP useful in field detection assays [2–6]. The use of LAMP in detection system has been explored for numerous micro-organisms and epidemic pathogen (Zika-virus, Ebolavirus, Malaria)[7] several of which validated by WHO [8], an FAO [2, 9]. SARS-CoV2 RT-LAMP tests were previously developed but to our knowledge, none of them propose a direct-from-swab, on field and all-in-one analytical solution such as ours [10–15].

### The innovation

We developed a specific RT-LAMP in dual format for the detection of SARS-CoV-2. The first one, named LoopXlab COVID-19 operates without extraction in conventional thermalcyclers (using 96 wells plates or 8 wells strips). The second one, named LoopXplore COVID-19, is designed for Point of Care (without extraction) and runs in the LoopX device (in 1.5 ml µtube). The objective of the study was to evaluate the performance of our two detection methods.

## MATERIAL & METHODS

### Study design, clinical samples and molecular tests

Nasopharyngeal samples (n=348) from patients with medical prescription were collected for virological diagnostic at the virology unit of CAEN University Hospital Center during April, May and June 2020. All samples were submitted to routine molecular diagnosis first and declared to be used in research.

Two available RT-qPCR systems were compared to our tests. The PCR developed by the french national reference center for respiratory viruses (targeting RdRp with IP2 / IP4 systems, and E gene) and a commercial multiplex real time RT-qPCR targeting the RdRp, E and N viral genes (Allplex 2019-nCov assay kit, Seegene, Korea). RT-qPCR assays were performed on Lighcycler 480 (Roche) and CFX (Biorad), two broadly available thermalcyclers.

*RT-LAMP assay* (*LoopXlab COVID-19*): 5 µL of sample were added to 20 µL RT-LAMP mix containing 1X Warmstart mastermix (NEB E1700L), DNA-primer mix composed 1,2µM FIP, 1,2µM BIP, 0,4µM F3, 0,4µM B3, 0,6µM LB and of a fluorescence dye (NEB E1700L). Negative controls (water) and a positive control (plasmid) was included in each experiment (Eurofins Genomic, Germany). The LoopXlab SARS-CoV2 was tested for 45 minutes runtime on Lightcycler 480 (Roche) – 63°C, Rotorgene (Qiagen) – 62°C, CFX (Biorad) – 63°C and MIC (Biomolecular systems) – 62°C (Figure 1A). Results are visualized as an amplification curve, where the time to threshold (TT) value, defined as the time required for the fluorescence to exceed a threshold, is an equivalent to the Ct or Cq values in real-time PCR (Figure 1B).

**Figure 1:**
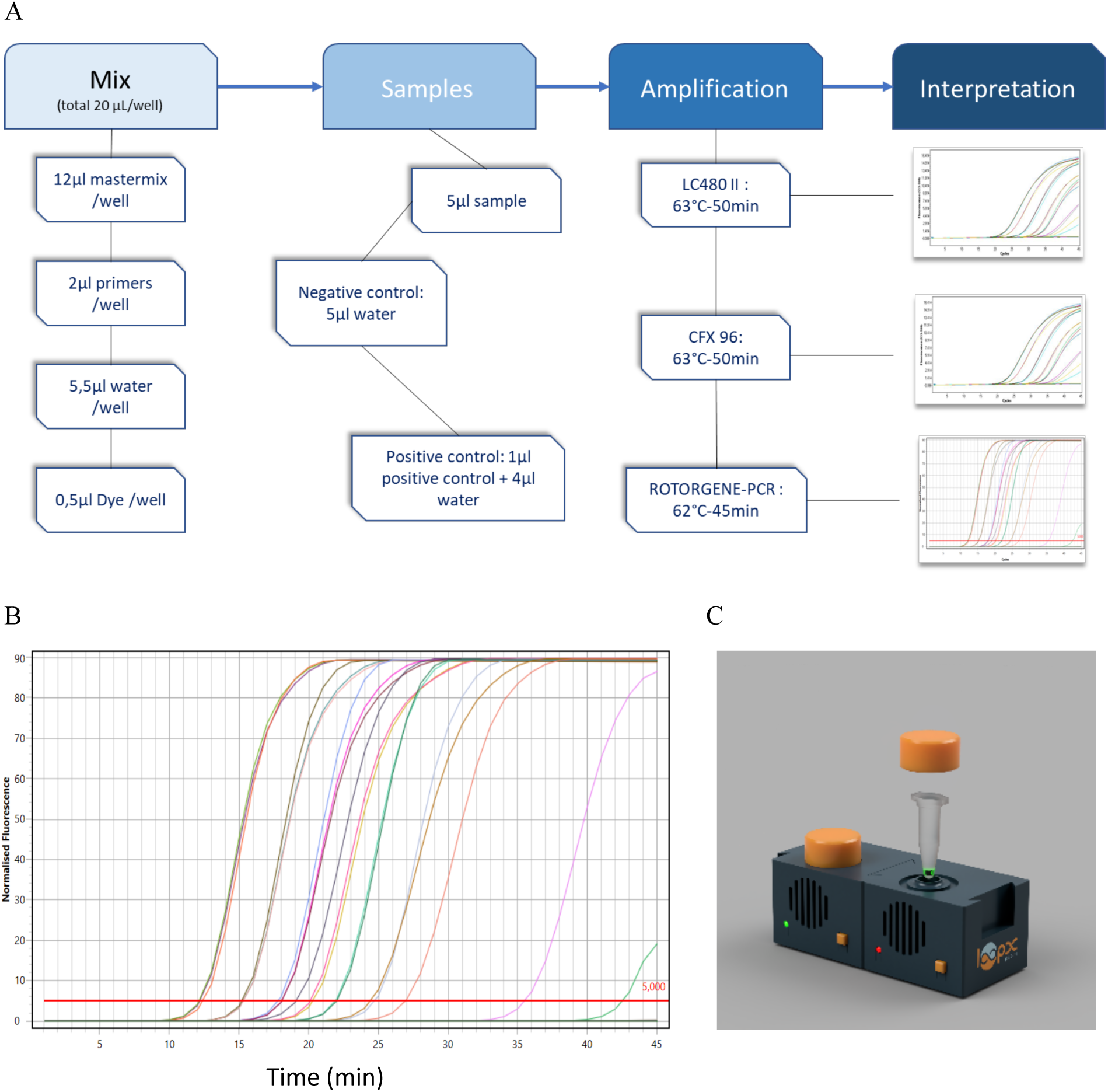
The LoopX test. A: LoopXlab COVID-19 flow chart, B: LoopXlab amplification curves, C: LoopX devices (2 paired units)

*RT-LAMP assay* (*LoopXplore COVID-19*): Twenty-five µL of sample were added to a microtube containing lyophilized mix including 20 µL RT-LAMP mix composed of 1X Warmstart mastermix (NEB M1702B), DNA-primer mix (0,8 µM FIP, 0,8 µM BIP, 0,25µM F3, 0,25µM B3, 0,4µM LB with specific quenching). RT-LAMP reaction and fluorescence detection were performed on LoopX (LoopDeeScience) device and last for 45 minutes at 63°C. LoopX device (Figure 1C) concentrates technologies allowing to heat reaction tube and read different wavelengths. Its dimensions (6×6×6 cm) and low energy requirements (USB-c connection) allow a simple Point Of Care analysis. The reaction tube carrying the sample is directly inserted into the LoopX chamber. The result is given directly without opening tube, thus preventing contamination. A specific program stored within the motherboard microprocessor controls the temperature and the wavelengths emission and reading (using diodes and spectrophotometer). An artificial intelligence calculates the Figure of Merit and give a false or true result displayed directly on the LoopX (red and green lights for positive and negative results, respectively) or on a bluetooth connected smartphone.

## RESULTS

### Linearity and limit of detection

The linearity for both RT-LAMP and RT-qPCR gave similar results (Figure 2). The limit of detection was calculated by limiting dilution. For each dilution point, RT-LAMP and RT-qPCR assays were tested in triplicates. For the latest dilutions (<100 RNA copies), the RT-qPCR Ct (Cycle threshold) exceeded 35. The LoopXplore test (POC) and the RT-qPCR gave similar results. In addition, Linearity and limit of detection were tested on CFX (Biorad), Lightcycler 480 (Roche), MIC (Biomolecular systems) and RotorGene (Qiagen) machines.

**Figure 2:**
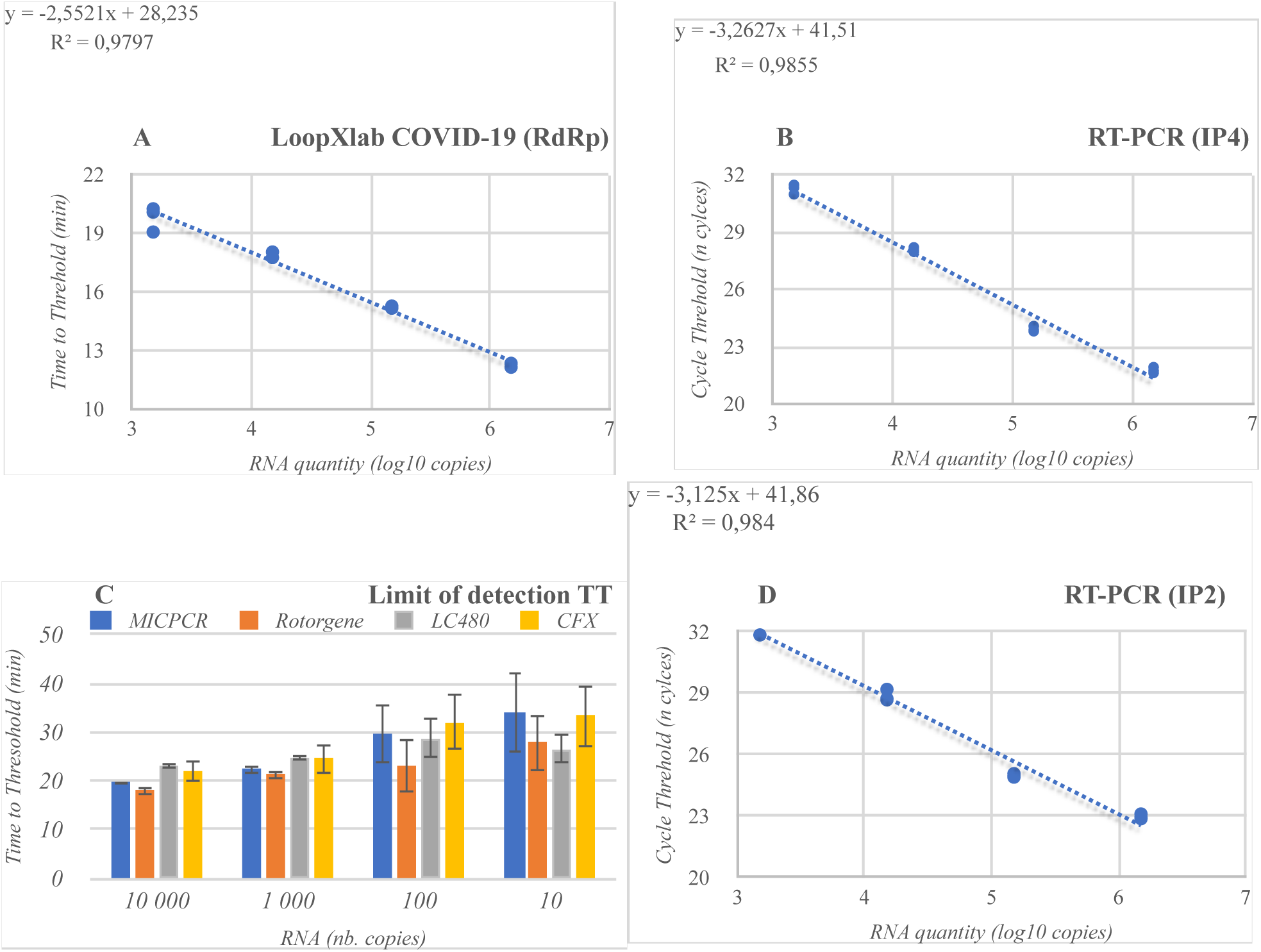
Linearity and limit of detection of LoopXlab and RT-PCR tests. A, B and D: linearity tests for LoopXlab and RT-PCR. Data were obtained using RNA standards extracted from the UCN1 SARS-CoV2 strain isolated and maintained in the virology unit of the University Hospital Center of Caen, Normandy, France. C: limit of detection for LoopXlab and stability of TT (time threshold) above 100 RNA copies.

### Specificity and sensitivity

LoopXlab and the LoopXplore POC tests were negative for a total of 52 respiratory samples positives for non-SARS-CoV2 coronaviruses, Rhino/Enteroviruses, Influenzavirus, RSV and other respiratory pathogens (Table 1). The validation consisted in testing 177 RNA extracts to optimize the RT-LAMP step and 171 crude samples without extraction to measure potential inhibition. Most widely used transport media were tested (Table 2). Media containing significant amount of EDTA or Ethanol (known inhibitors for the LAMP reaction) inhibit the reaction and cannot be used in our tests without extraction (Table 2). All samples were analyzed twice by RT-qPCR and both RT-LAMP technics (LoopXplore in LoopX and LoopXlab in thermalcycler). RT-LAMP methods have similar sensitivity and specificity (LoopXlab: 98.6% and 91.5%, LoopXplore: 95.2% and 95.5%, respectively) Only one sample was not detected by loopXlab (RT-qPCR CT value of 32) and LoopXplore (CT >35)(Table 3).

**Table 1:** Selectivity essay for LoopXlab and LoopXplore performed on lighcycler 480 and LoopX respectively.

| **Samples**<br>(machine) | **N tests** | **LoopXlab**<br>(LC480) | **LoopXplore**<br>(LoopX) |
| --- | --- | --- | --- |
| **SARS-COV2** | 2 | **2/2** | **2/2** |
| **Coronavirus HKU1** | 6 | 0/6 | 0/6 |
| **Coronavirus NL63** | 1 | 0/1 | 0/1 |
| **Coronavirus OC43** | 1 | 0/1 | 0/1 |
| **Adenovirus** | 7 | 0/7 | 0/7 |
| **Bocavirus** | 3 | 0/3 | 0/3 |
| **Chlamydia pneumophila** | 1 | 0/1 | 0/1 |
| **Entéro / Rhinovirus** | 13 | 0/13 | 0/13 |
| **Influenza A H1N1v2009** | 2 | 0/2 | 0/2 |
| **Influenza A H3N2** | 1 | 0/1 | 0/1 |
| **Influenza B** | 4 | 0/4 | 0/4 |
| **hMPV** | 4 | 0/4 | 0/4 |
| **VRSA** | 8 | 0/8 | 0/8 |
| **VRSB** | 1 | 0/1 | 0/1 |

**Table 2:** List of transport media tested with LoopXlab technology. “Compatible media” indicates the absence of inhibition. “Not optimal” indicates that inhibition is rarely observed. “Not compatible” indicates that inhibition is systematically observed. For media not listed here, inhibition controls are recommended.

| Name of the media | Supplier | Remarks and comments |
| --- | --- | --- |
| Physiological water | - | Compatible |
| Σ-virocult® | MWE | Compatible |
| MSwab® | Copan | Compatible |
| MEM | Gibco | Compatible |
| Σ-VCM® | MWE | Not optimal |
| UTM™ | Copan | Not optimal |
| UTM™ | Noble Biosciences | Not optimal |
| Cytoswab | WellKang | Not optimal |
| Yocon® Viral Transport Media | Yocon Biology Technology Company | Not compatible |
| Nest® ITM | Wuxi Nest Biotechnology Co., Ltd | Not compatible |
| Cobas® PCR Media | Roche Diagnostics GmnH | Not compatible |

**Table 3:** Sensitivity and specificity for LoopXlab and LoopXplore with and without extraction. qRT-PCR ct values were given by the IP2-IP4 system from national reference lab.

| Methods | LoopXLab |  | LoopXPlore |  |
| --- | --- | --- | --- | --- |
| Sample matrix | extracted RNA | Specimen<br>(no extraction) | extracted RNA | Specimen<br>(no extraction) |
|  | N positive qRT-PCR / N positive LoopX |  |  |  |
| ct < 20 | 7 / 7 | 7 / 7 | 8 / 8 | 6 / 6 |
| 20ct ≤ ct < 30ct | 15 / 15 | 33 / 33 | 6 / 6 | 7 / 7 |
| ct ≥ 30 | 4 / 3 | 29 / 28 | 4 / 4 | 8 / 7 |
| No ct | 95 / 95 | 59 / 54 | 38 / 38 | 22 / 21 |
| Sensitivity | 96,2% | 98,6% | 100,0% | 95,2% |
| Specificity | 100,0% | 91,5% | 100,0% | 95,5% |

## CONCLUSIONS & RECOMMENDATIONS

LoopXlab COVID-19 detection kit test was designed in the context of covid-19 early response and came with a dedicated all-in-one device ensuring automated, reproducible and optimal reaction and reading. The sensitivity and the specificity were of 98.6% and 91.5% respectively. This convenient extraction-free protocol drops the sample-to-result time below one hour (45 minutes of cycling). This test provides a more efficient while sensitive and reliable SARS-CoV2 molecular testing and stands for a serious alternative to conventional RT-qPCR tests in either massive parallel testing or point-of-care formats. The massive parallel format would greatly increase testing capacities of diagnostic labs while the Point-of-Care format is very easy and accessible to non experts. Both formats are very safe as the tube remain sealed after sample addition and reading is automated. Adaptation of these tests for saliva testing is currently under evaluation.

## Data Availability

Data used in the present study are available upon request.

## ACKNOWLEDGMENTS

The authors wish to acknowledge the Team from New England Biolabs France for technical support, the Virology unit and all contributors to the study of University Hospital of Caen and Loo Dee Science company, the “Agence de l’Innovation de Défense” (French Defence Innovation Agency) and the “DGA Maitrise NRBC” (Armament General Directorate CBRN Defence Center).

